# Repurposing EEG monitoring of general anaesthesia for building biomarkers of brain ageing: An exploratory study

**DOI:** 10.1101/2022.05.05.22274610

**Authors:** David Sabbagh, Jérôme Cartailler, Cyril Touchard, Jona Joachim, Alexandre Mebazaa, Fabrice Vallée, Étienne Gayat, Alexandre Gramfort, Denis A. Engemann

## Abstract

**Background:** EEG is a common tool for monitoring anaesthetic depth but is rarely reused at large for biomedical research. This study sets out to explore repurposing of EEG during anaesthesia to learn biomarkers of brain ageing in the absence of consciousness.

**Methods:** We focused on brain age estimation as an example. Using machine learning, we reanalysed 4-electrodes EEG of 323 patients under propofol and sevoflurane. We included spatio-spectral features from stable anaesthesia for EEG-based age prediction applying recently published reference methods. Anaesthesia was considered stable when 95% of the total power was below a frequency between 8Hz and 13Hz.

**Results:** We considered moderate-risk patients (ASA <= 2) with propofol anaesthesia to explore predictive EEG signatures. Average alpha-band power (8-13Hz) was informative about age. Yet, state-of-the-art prediction performance was achieved by analysing the entire power spectrum from all electrodes (MAE = 8.2y, R2 = 0.65). Clinical exploration revealed that brain age was systematically linked with intra-operative burst suppression – commonly associated with age-related postoperative cognitive issues. Surprisingly, the brain age was negatively correlated with burst suppression in high-risk patients (ASA = 3), pointing at unknown confounding effects. Secondary analyses revealed that brain-age EEG signatures were specific to propofol anaesthesia, reflected by limited prediction performance under sevoflurane and poor cross-drug generalisation.

**Conclusions:** EEG from general anaesthesia may enable state-of-the-art brain age prediction. Yet, differences between anaesthetic drugs can impact the effectiveness of repurposing EEG from anaesthesia. To unleash the dormant potential of repurposing EEG-monitoring for clinical and health research, in the absence of consciousness, collecting larger datasets with precisely documented drug dosage will be key enabling factors.

## Introduction

Electroencephalography (EEG) is often used during general anaesthesia (GA) for monitoring anaesthetic depth and adjusting drug dosage^1–5^. The increasing availability of EEG recordings from GA has stimulated cross-cutting research linking anesthesiology, neuroscience and cognition^6–8^. EEG dynamics during GA and the drug dosage required for achieving stable anaesthesia can depend on age and general health^9^. Cognitive decline has been associated with reduced drug requirement and EEG-power changes in the alpha frequency band (8-13Hz) ^10,11^. These recent results point out the possibility that individual brain health can be reflected in EEG signals observed during GA^12^.

Could EEG during GA, thus, be used beyond anaesthesia monitoring as a screening tool to assess the risk of developing neurodegenerative diseases? Several factors are favourable to this idea. First, EEG signals during stable anaesthesia can be of high signal quality as administration of curare minimises movement artefacts. Second, the availability of EEG recordings from GA keeps growing as GA is conducted in all age groups and everywhere in society, potentially avoiding selection bias of laboratory research^13,14^. Third, recent progress in advanced analytics and machine learning for EEG has led to development of novel biomarkers of cognition, brain function and health^15–17^. When combined, these factors could turn EEG monitoring data from GA into a valuable tool for clinical research.

In this study, we explore the possibility of repurposing existing EEG monitoring data from GA for building data-driven measures of brain ageing^18^. Concretely, we propose to translate the previously developed brain-age framework^19^ to the context of GA. By using machine learning, brain age can capture individual ageing. This framework provides quantitative comparisons of the chronological age of a person against their statistically expected age given their brain data (“this brain looks older/younger”)^20,21^. Over the past years, numerous studies^19,20,22–24^ have found that brain age estimates from the general population yield sensitive measures of neurodegenerative risk and disease severity in clinical populations^25^.

The bulk of the brain age literature is based on MRI^19,24,26,27^. Nevertheless, a recent line of research has demonstrated that brain age can be estimated from EEG and may even contribute additional information over MRI^20,28,29^. It is currently unknown if brain age can be estimated from EEG during GA. Most EEG approaches for brain-age modelling relied on fine-grained spatial information provided by dozens to hundreds of electrodes and data collected in the laboratory setting^17,28^. On the other hand, brain age has recently been estimated from sleep EEG with fewer electrodes^18,30^. Another source of potential complications for estimating brain age from EEG during GA is related to the type of anaesthetic drug. For example, compared to widely used propofol anaesthesia, halogenated gases can shape EEG activity differently, increasing modelling complexity. Moreover, the amount of anaesthetic drug given to a patient to achieve a stable anaesthesia may by itself reflect the patient’s cognitive health^10^, potentially inducing confounding effects.

Our primary objective was to demonstrate the feasibility of modelling chronological age as a function of EEG power from intraoperative recordings in moderate-risk patients (ASA <= 2). We used state-of-the-art machine learning methods to estimate model prediction performance and hypothesised that the entire power spectrum was informative rather than specific frequency bands commonly used for monitoring anaesthetic depth. Our secondary objective was to investigate the association between chronological age, EEG-derived brain age, and intraoperative burst suppression in moderate-risk patients (ASA <= 2) and those suffering from pronounced health risk (ASA = 3). Finally, we investigated the specificity of the propofol-induced EEG signature of brain age through generalisation testing against EEG collected under sevoflurane anaesthesia.

## Methods

We reanalysed EEG and clinical data from the Probrain study registered under the ID NCT03876379 and approved by the SRLF (Société de réanimation de langue française) Ethics Advisory Committee (Chairperson Dr Jean Reignier, 48, avenue Claude Vellefaux, Paris, France) on the 5 January 2016, under the reference CE SRLF 11-356. The SRLF is the French national academic society for anaesthesia and critical care consulted by the department of anaesthesiology at the Lariboisière hospital (Paris, France). Patients were provided with an information letter. Verbal consent was recorded from every patient before anaesthesia.

### Patient selection

Between September 2017 and January 2020, patients eligible for an elective interventional surgery or interventional procedure under GA (for orthopaedic surgery or neuroradiology intervention for asymptomatic aneurysm) were selected to participate in the prospective, observational, monocentric Probrain study at the Lariboisiere hospital. The following exclusion criteria applied: pregnant women, age below 18 years, patients on sedation and mechanical ventilation before the procedure, history of bleeding aneurysm, neurodegenerative disease, neurological disorders, untreated depression.

### Anaesthetic protocol

GA was carried out based on standard practices. An opioid was administered (0.2ug/kg/h of sufentanil for orthopaedics and 3-5ng/ml of remifentanil for neuroradiology) followed by intravenous propofol and atracurium besylate non-depolarizing curare for GA induction. During maintenance either intravenous propofol or sevoflurane vapors were used as the main anaesthetic agents. Propofol was administered using total intravenous anaesthesia with a brain effect site concentration ranging from 3 to 3.5 ug/ml set by the physician and maintained automatically by syringe drivers under the Schnider model^31^. For sevoflurane, the mean alveolar concentration was set to obtain exhaled concentration at 1.5 to 2%. The choice of the anaesthetic drug for GA maintenance between sevoflurane and propofol, was left to the discretion of the anesthesiologist in charge. Propofol TCI (Target Controlled Infusion) and sevoflurane MAC (Minimum Alveolar Concentration) were adapted to keep a SEF95 between 8 and 13Hz during the maintenance.

The mean arterial pressure during the entire intervention was maintained at 90% of its reference value and always above 65mmHg.

### Data collection

Cerebral activity during GA was monitored using a Masimo^™^device with a 4-frontal electrodes EEG montage (Fp1, Fp2, F7 and F8, referenced on Cz), sampled at 63 Hz by default. EEG sub-hairline electrodes were placed a few minutes before GA induction and removed shortly after recovery of consciousness. Intraoperative EEG data were then extracted from the device, anonymized and stored on a file server in EDF format. We collected demographic data (age, gender, weight, height, BMI), clinical information (type of anaesthetic drug, ASA score) from the anaesthesia-specific medical consultation. This resulted in n = 323 patients with both a proper concatenated EEG recording and metadata information. The resulting sample is described in Table 1.

**Table 1.**
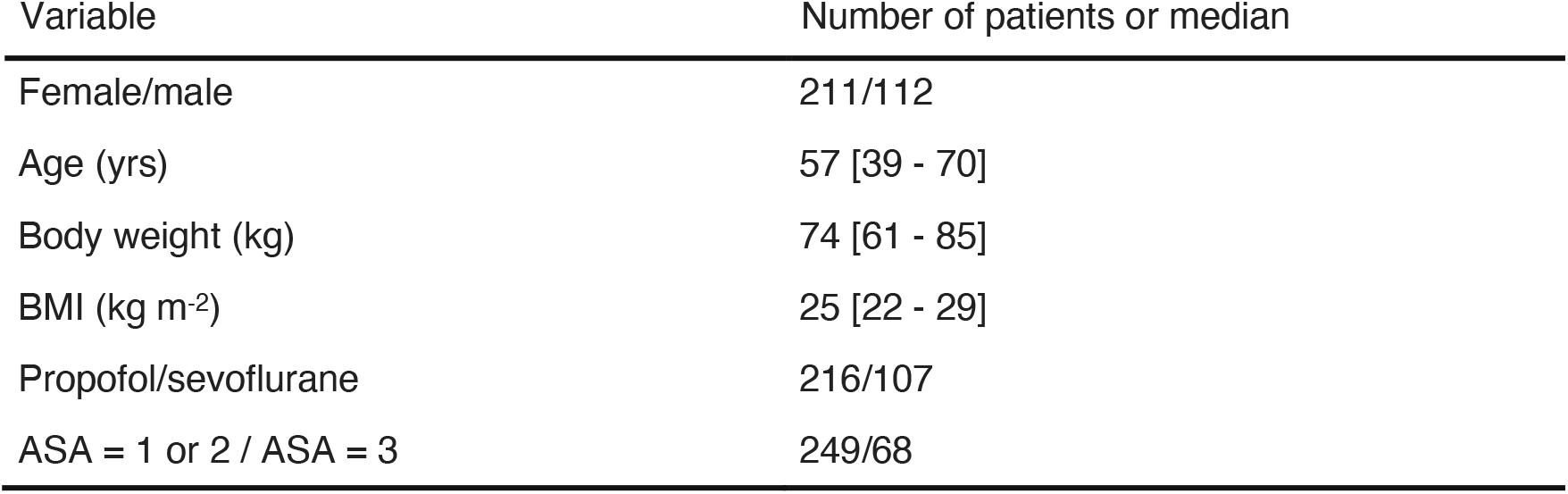
Patient characteristics. Data are presented as median [25^th^–75^th^ percentiles].

### EEG processing, model construction and clinical exploration

The EEG processing and feature extraction is outlined in Fig 1 and detailed in the supplementary materials. We applied brain age models and methods for model comparisons from previous reference publications^20,28^. Ridge regression was used as prediction algorithm with 100 (Monte Carlo) cross-validation (CV) splits. Model superiority was assessed via pair-wise ranking across CV splits^20,32^. For model construction, we focused on moderate-risk patients with low ASA scores under propofol anaesthesia (strictly below 3, n = 170, see ds1 in **Table 2**). For clinical exploration, we compared brain age to burst suppression (a common predictor of postoperative complications). We computed cross-validated brain age predictions^20^ on the data used for model building (ASA <= 2, ds3 in **Table 2**) and then predicted brain age in a dataset from a high-risk population not used for model building (ASA = 3, ds4 in **Table 2**). The brain age variable was then obtained by concatenating the predictions across the two datasets (ds5 in **Table2**). For definition of burst suppression and details on statistical analyses and software, see supplementary materials.

**Table 2.** Subsets of patients used for model construction and model exploration defined by ASA status and drug type (total including patients without burst suppression annotation)

| Subset | Number of patients |
| --- | --- |
| ds1 [construction]: ASA ≤ 2, propofol | 170 |
| ds2 [exploration]: ASA ≤ 2, sevoflurane | 79 |
| ds3 [exploration]: ASA ≤ 2, propofol | 167 (170) |
| ds4 [exploration]: ASA = 3, propofol | 37 (40) |
| ds5 [exploration]: ASA ≤ 3, propofol | 204 (210) |

**Fig 1.**
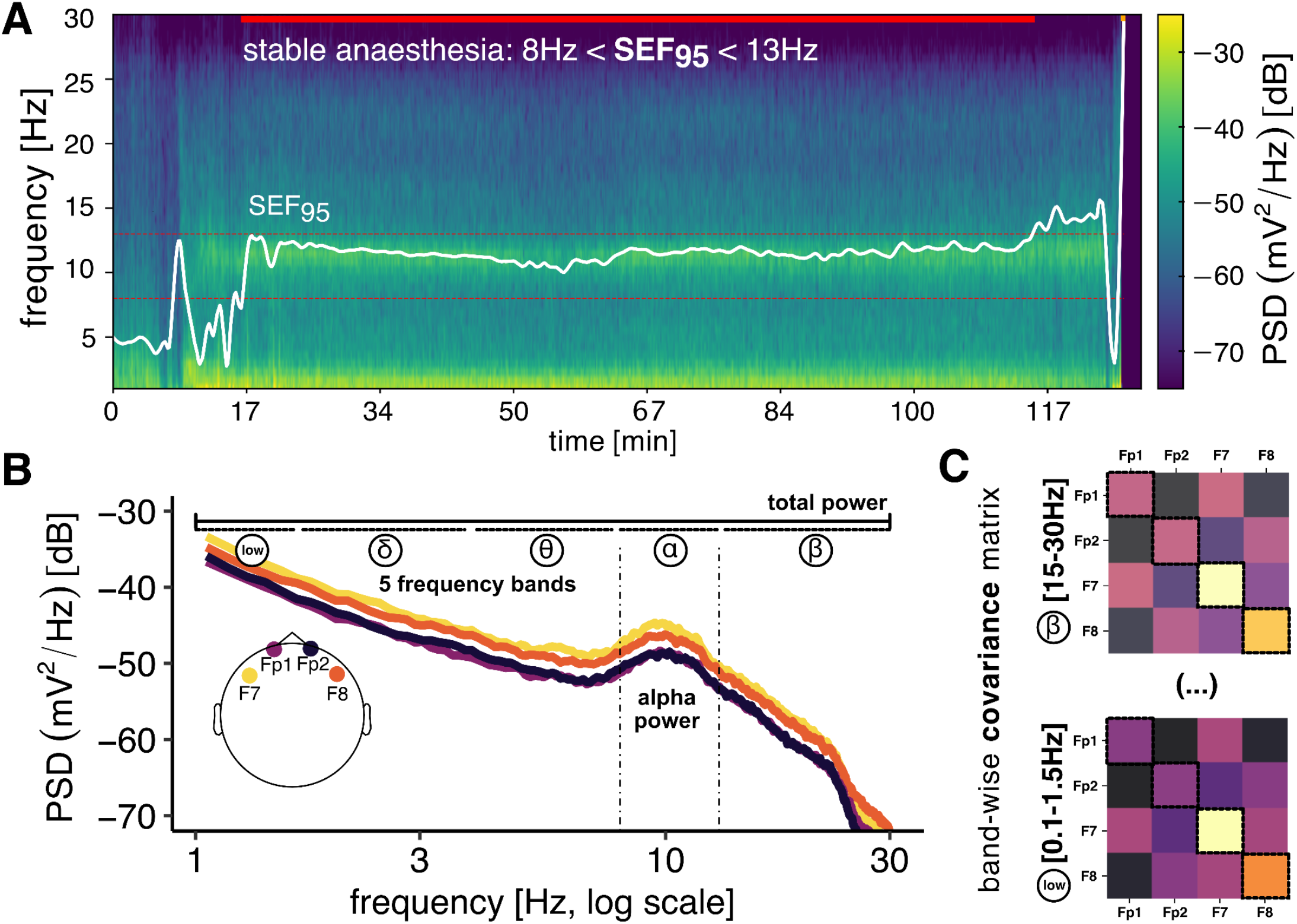
Building biomarkers of brain aging from stable segments of GA. **(A)** We selected the longest consecutive period of stable anaesthesia (propofol), defined by the spectral edge frequency (SEF) 95 falling in the 8-13 Hz range, the absence of high-voltage artifacts and of burst suppression. **(B)** We estimated the power-spectral density (PSD), which can reveal EEG signatures commonly used in anaesthetic monitoring such as total power or alpha power. Furthermore, we investigated the hypothesis that additional information on targets beyond clinical monitoring, *i*.*e*., brain age is distributed across the entire power spectrum. For this purpose, we adaptively summarized the PSD over all frequencies and channels using supervised learning. Reference models for estimating brain age from EEG have exploited spatial patterns in different frequency bands presented by the covariance between EEG channels **(C)** whose diagonal summarises the PSD in the given frequency band. To assess the complementarity of these readouts, we used the stacking approach in which 1) ‘alpha power’, 2) ‘total power’, 3) the ‘power spectrum’ and 4) ‘spatial patterns’ from 5 frequency bands were gradually included. Model comparisons established the relative merit of these increasingly complex features.

## Results

We considered 323 patients for which we obtained actionable EEG (**Table 1**). A flow chart summarising the data collection process is shown in Supplementary Materials (**Fig S1**). Analysis-defining subsets of patients are listed in **Table 2**. We first explored the relationship between EEG activity and ageing across the frequency spectrum under propofol anaesthesia (**Fig 2**), which was more commonly administered in this study (**Table 1, Table2:** ds1). Binning the power spectra by age groups revealed age-related patterns (**Fig 2A**). Across frequencies, younger patients tended to show higher EEG power. We formally quantified this tendency using a linear mixed-effects model regressing the log power (dB) on age, log frequency and their interaction (intercepts varying by patient). The analysis uncovered that regardless of frequency, EEG power declined on average by -0.10 dB, 95% CI [-0.13, -0.08] with every year of age. The full model is summarised in the supplementary materials (**Table S1**).

**Fig 2.**
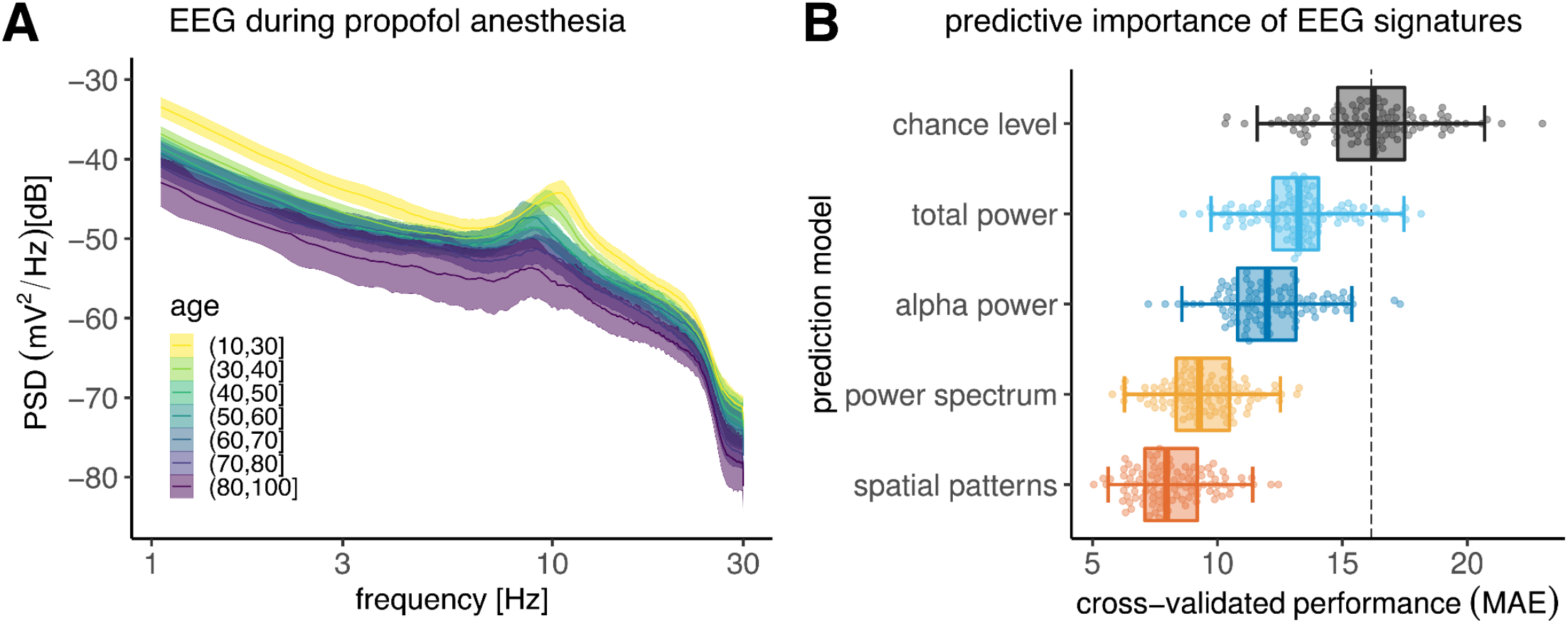
Modelling brain ageing from spatio-spectral EEG signatures during propofol anaesthesia. **(A)** depicts average power spectral density (PSD) by age group represented by different colours (n = 170, ASA <= 2). Shaded areas represent twice the standard error of the mean for every age group. Power and peak frequencies tend to decrease with age. **(B)** depicts cross-validated performance of age-prediction models based on progressive inclusion of different EEG signatures (Monte Carlo, 100 splits, MAE = mean absolute error). Dots represent scores at individual cross-validation splits, boxplots summarise the cross validation distribution, whiskers show the 2.5^th^ and 97.5^th^ percentiles. The dotted line indicates the expected performance for a chance-level model. Results are ordered by prediction error, descending from top to bottom. Models include EEG signatures from previous models (e.g. ‘alpha power’ refers to combining total power and alpha power). Informal analyses revealed that spatial patterns enabled virtually identical performance as the final model combining the diverse EEG signatures. For simplicity, we focused on the spatial patterns model in all subsequent analysis.

**Fig 2B** presents model comparisons for combinations of different EEG signatures. Chance level was estimated around 16 years of mean absolute error (MAE; P_25_ = 14.8, P_75_ = 17.5). The ‘total power’ model led to an improvement of the cross-validation (CV) score by about -2.9 years MAE on average (P_25_ = -4.12, P_75_ = -1.9), performing better than chance in 95/100 CV splits. As average alpha power was included, the CV scores improved by another -1.2 years MAE (P_25_ = -1.9, P_75_ = -0.34), beating the previous model on 86/100 CV splits. Adding fine-grained spectral information across all frequencies from 1-30Hz led to further improvements by -2.7 years MAE (P_25_ = -3.7, P_75_ = -1.86), out-performing the previous model on 95/100 CV splits. Finally, including information about the spatial patterns of covariance between the electrodes lowered the CV score by another -1.22 years MAE (P_25_ = -1.93, P_75_ = -0.47 superiority: 85/100 CV splits). In absolute terms, the final performance was about 8.2 years MAE (P_25_ = 7.09, P_75_ = 9.19) and corresponded to an R^2^ score equivalent to 65% of explained variance (P_25_ = 0.58, P_75_ = 0.75).

For clinical exploration, we focused on the age-prediction signal under propofol anaesthesia in a subset of patients for whom burst suppression was annotated (n = 204, **Table 2:** ds3-ds5). We investigated the link between brain age, health status and undesirable intraoperative burst suppression. **Fig 3A** plots model-based age predictions against chronological age and burst-suppression proportion for the previously analysed cohort of patients with good general health (ASA score <= 2; n = 167, **Table 2:** ds3) as well as extrapolations to a distinct cohort of patients (ASA score = 3; n = 37, **Table 2:** ds4). Results suggested more complex relationships between EEG-predicted age and burst suppression across patients of different health status and chronological age: One can observe that in younger patients with ASA <= 2, burst suppression was more frequent among those with higher brain age. This trend seems inverted in older patients with ASA = 3 where bigger dots concentrate under the diagonal identity line. To formalise these relationships, we modelled the logit of the proportion of burst suppression as a weighted sum of scaled age, scaled brain age and health status and their respective interaction terms (**Fig 3B**). The analyses revealed a significant effect of brain age (beta = 0.85, SE = 0.20, t(196) = 4.4, p < 0.0001). Given a standard deviation of brain age of about 16 years, this suggests that, across age and health status, burst suppression increased by 134% (exp(0.85) = 2.34) for every 16 years of brain age. Furthermore, a significant effect of brain age and health status emerged (beta = -0.99, SE = 0.44, t(196) = -3.7, p = 0.03), suggesting that the proportion of burst suppression was reduced by 63% (exp(−0.99) = 37%) for every 16 years of brain age in patients with ASA = 3 as compared to the other patients across all chronological ages. The full model is reported in the supplementary materials (**Table S2**).

**Fig 3.**
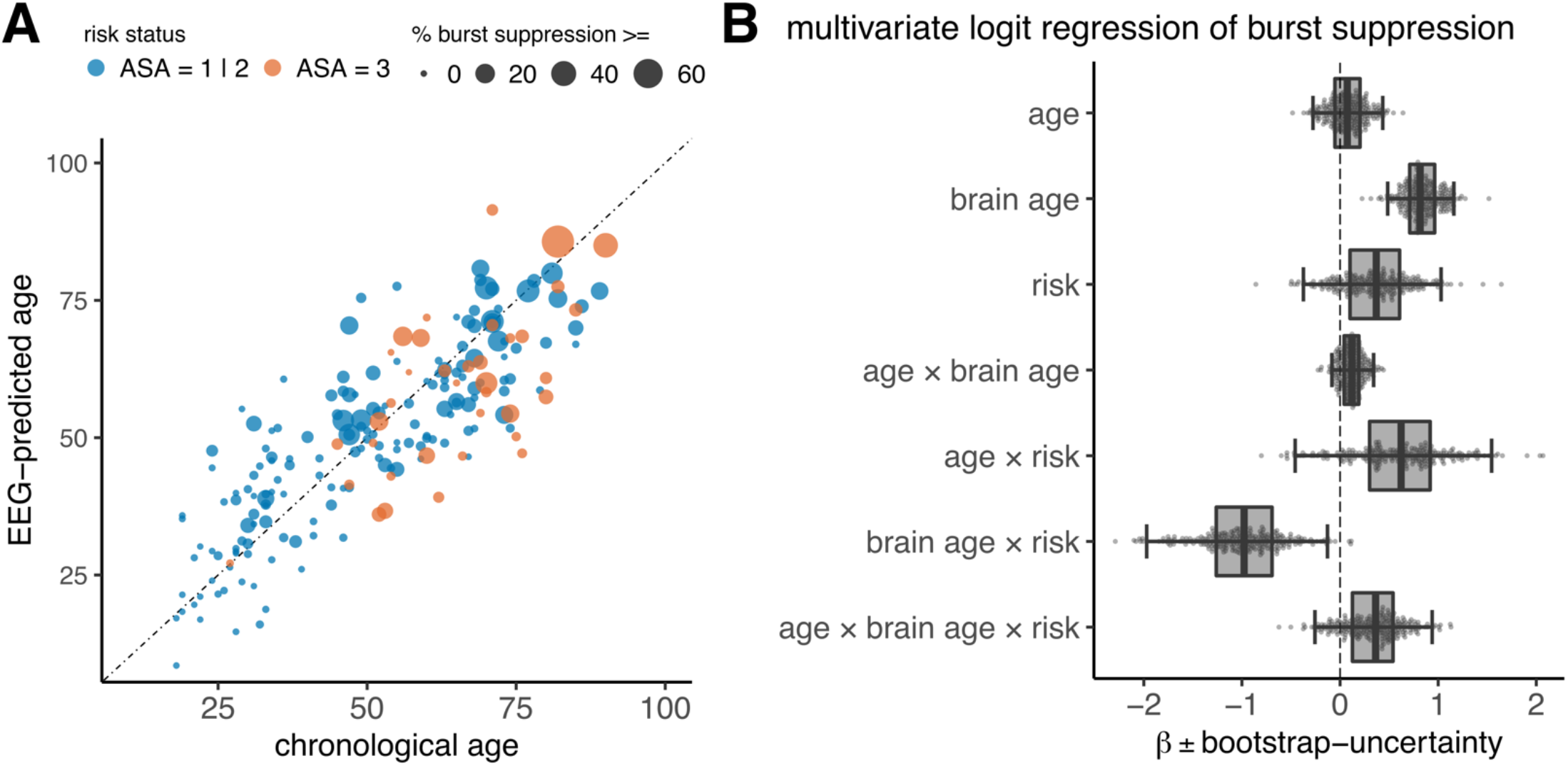
Clinical exploration of Brain age derived from EEG during anaesthesia. **(A)** plots chronological age (x axis) and brain age (y axis) against the proportion of the EEG showing burst suppression (marker size). Binary risk status (according to ASA scores) isi indicated by colour (n = 204). **(B)** presents a logit-regression model summarising these trends. Box plots represent the two-sided 95% CI (quantiles 2.5, 97.5), individual dots indicate possible coefficients implied by the statistical uncertainty of the model. Brain age and the interaction with brain age showed most consistent effects. Note that the exponential values of the coefficients can be interpreted as odds ratios of the proportion of burst suppression.

Visual inspection of EEG activity under sevoflurane anaesthesia (**Fig 4A, Table2:** ds2) suggested higher EEG-power levels and weaker correlation between EEG power and age. We formally contrasted these trends with the previous results (**Fig 4A**) through a linear mixed-effects model regressing the log power (dB) on age, log frequency, drug type and their interactions; intercepts varied by patient. Independently of frequency or drug type, EEG power declined by - 0.10 dB, 95% CI [-0.13, -0.08] for every year of age. Compared to propofol, sevoflurane led to 3.6 dB higher EEG power, CI 95% [1.21,6.10]. Notable interaction terms pointed at more complex EEG patterns: First, a two-way interaction suggested that differences between propofol and sevoflurane depended on frequency, implying that, under sevoflurane, the power declined, on average, by about -1.00 dB more than under propofol per Hz, 95% CI [-1.27, -0.76]. Second, a three-way interaction suggested that under sevoflurane the effect of age may non-linearly increase log power by 0.01 dB per Hz, 95% CI [<0.01,0.01]. The full model is reported in the supplement materials (**Table S3**).

**Fig 4.**
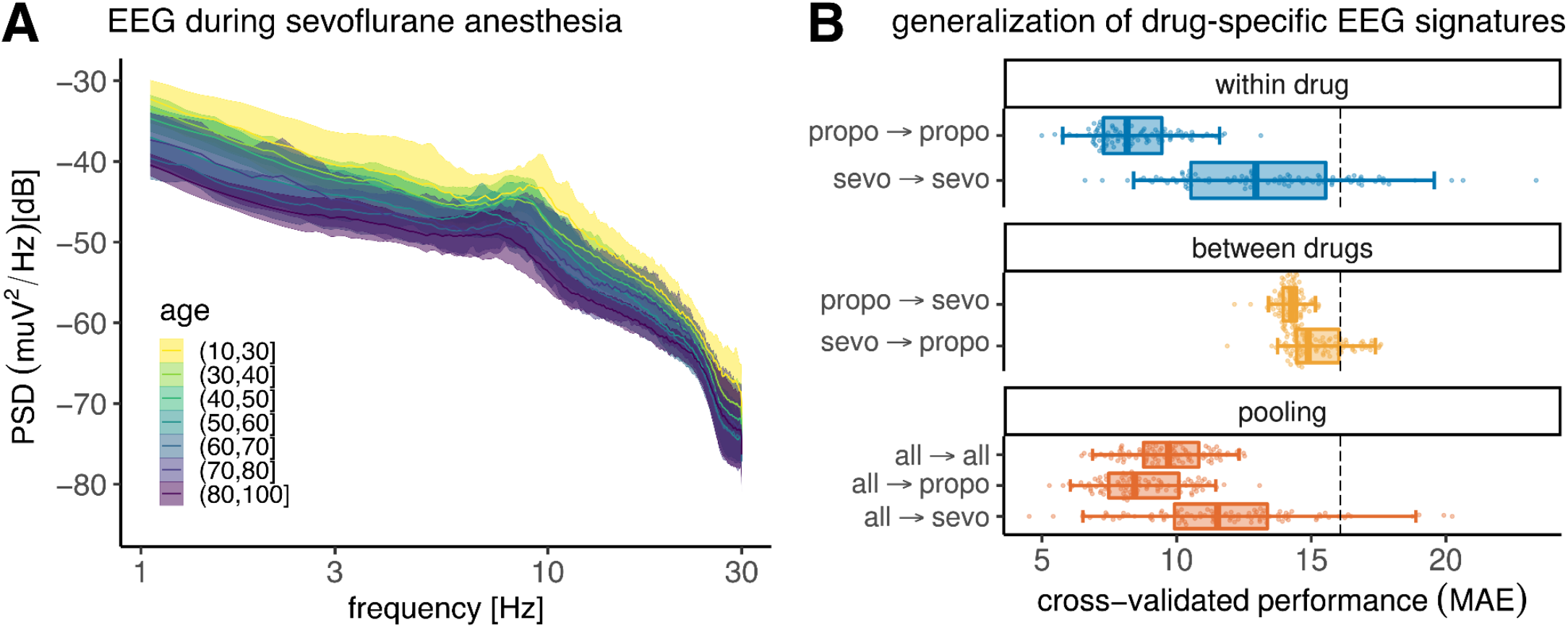
Divergence and pooling of EEG-signatures by drug type. **(A)** depicts the power spectral density of EEG under sevoflurane anaesthesia (n= 79, ASA <= 2). Visual conventions as in Fig 1A. Age appeared less consistently revealed by the PSD from sevoflurane as compared to propofol anaesthesia, raising the question if age prediction can be compared between these two drugs. **(B)** breaks down modelling performance of the ‘spatial patterns’ model by drug type (100 Monte Carlo splits, conventions as in Fig 1B). Within-drug models were fitted and evaluated separately by drug type (blue). Between-drug models were fitted on one drug type but evaluated on the other drug type (orange). Pooling was attempted (dark orange) by combining data for fitting and evaluation on both drug types (first row). Separate scoring was performed by drug type (rows 2-3). Results suggest that performance was best when using EEGs from propofol anaesthesia. While generalisation between drug types was poor, pooling EEGs from different drug types preserved the individual drug-specific performance.

**Fig 4B** presents comparisons between age-prediction models that were either separately fitted and evaluated within each drug type, fitted within one drug type and evaluated on the other drug type, or fitted and evaluated pooling both drug types. When fitted separately, the prediction model from sevoflurane EEG performed about 4.9 years of MAE (P_25_ = 2.7, P_75_ *=* 7.0) worse than propofol, inferior on 95/100 CV splits. Similarly, cross-drug generalisation of prediction models led to low prediction performance between 14 and 15 years of MAE. By comparison, generalisation from propofol to sevoflurane EEG was by -1 year of MAE more successful (P_25_ = -1.8, P_75_ = -0.25, superiority: 86/100 CV splits). Prediction models trained on the pooled EEG data from both drug types led to worse performance than the model trained under propofol EEG with about 1.3 year higher MAE (P_25_ = -0.31, P_75_ = 2.8, inferiority: 68/100 CV splits). On the other hand, results improved over the sevoflurane-based model by -3.7 years of MAE (P_25_ = -6.4, P_75_ = -1.3, superiority: 85/100 CV splits). Considering the subscores of the pooled model for the observations under sevoflurane EEG, suggested that combining both drugs led to an improvement of -1.6 years of MAE (P_25_=-1.01, P_75_= 4.0) compared to the model trained on sevoflurane EEG (superiority: 67/100 CV splits)

## Discussion

This study explored the feasibility of repurposing EEG-monitoring data from GA for building measures of brain ageing by importing machine learning approaches for brain age prediction originally developed in the laboratory setting. Under propofol anaesthesia, we reached prediction performance comparable to reference studies using research-grade EEG^28,33^. Model comparisons revealed that age-related information was present in spatial activity patterns distributed across the entire power spectrum. Clinical exploration of the well-performing propofol model highlighted important associations between brain-predicted age and the probability of burst suppression, which, however, depended on health status. Additional results suggested that the brain age signature was specific to propofol and may not generalise to sevoflurane.

Clinical studies have identified a link between EEG power spectra in the alpha band (8-13Hz), pre-operative cognitive decline^10^, intraoperative burst suppression^34,35^ and postoperative cognitive disorders^36^. This body of work pointed at the possibility that the EEG response to anaesthetic drugs may reveal the presence of neurodegenerative risk. Here we extended this prior art by directly applying brain-age prediction models for EEG^28^ on a larger dataset of EEG during anaesthesia. Under propofol, this approach has led to performance matching recent work with high-density MEG and EEG^20,37^. Previous studies during anaesthesia have instead focussed on EEG-signatures closely related to anaesthesia monitoring, with particular emphasis on total power and alpha power^6,38^. Our results have shown that the entire power spectrum and fine-grained correlations between signals collected at different electrodes may contain information relevant for learning biomarkers beyond anaesthesia monitoring.

On the other hand, prediction models constructed from EEG under sevoflurane anaesthesia were far less convincing and did not combine well with EEG collected under propofol anaesthesia. This may be intrinsically related to differences between both drugs regarding the mechanism of action: Propofol selectively activates GABA_A_ receptors^39^ whereas sevoflurane acts on several synaptic pathways, potentially increasing the complexity of the signal across age^40^. An alternative explanation might be that the dosage of sevoflurane was more constant across patients as it relied on standard MAC target values. Instead, propofol dosage was determined by a personalised TCI target value, chosen to stabilise GA i.e. to keep SEF95 in 8-13 Hz. This could, in principle, open the door to confounding effects, if propofol requirement for stable anaesthesia depends on age and health: Higher propofol dosage and brain age both dampen EEG power while patients at risk of developing burst suppression might receive lower doses of propofol. In turn, this should lead to reduced propofol-induced dampening, cancelling out the effect of brain-age on EEG power. That would be in line with observed changes of direction of association between brain age and burst suppression when comparing moderate-risk (ASA <= 2) versus high-risk (ASA = 3) patients, for whom, higher brain age went along with less burst suppression. Unfortunately, this hypothesis cannot be readily disambiguated using the present study, as neither the propofol dosage nor TCI parameters at a given moment in the EEG recording were available for deconfounding analysis.

The present study has successfully applied concepts and methods from laboratory research in cognitive neuroscience^19,23^ on EEG monitoring data collected during GA. Therein, our systematic model comparisons of EEG signatures, drug types and patient populations have extended the scope of brain-age research to clinical real-world EEG collected in the absence of consciousness. Our findings motivate future research in clinical and cognitive neuroscience in anesthesiology beyond monitoring of anaesthesia depth. These strengths of our work have to be put in perspective with the important limitation that inconsistent findings regarding the variability of brain-age effects across drug types could not be resolved. Despite successful age-prediction results, the current work, therefore, does not yet present a ready-to-use brain age measure as, for example, the MRI-based brain age delta^19^. To push this exploratory effort to the next level, future studies with, ideally, larger samples must focus on precise control and measurement of drug dosage at any moment during anaesthesia. Preferably, propofol should be administered at constant dosage to rule out confounding by clinical factors. Validation against pre-surgical brain age estimates derived from gold-standard anatomical MRI^24^ or research-grade high-density EEG^28^ will be essential.

In conclusion, the present study points out the general feasibility of repurposing EEG from anaesthesia for learning biomarkers of brain ageing and health beyond the imminent perimeter of patient monitoring. To unleash this dormant potential of EEG-monitoring data for clinical and public health research, collecting larger datasets with precisely documented drug dosage will be key enabling factors.

## Data Availability

All data produced in the present study are available upon reasonable request to the authors.

## Author contributions

authors in alphabetical order

**Conceptualization**: A.M., A.G, D.E., E.G., F.V.

**Data curation**: C.T., D.S., J.J., F.V.

**Software**: A.G., D.E., D.S., J.C.

**Formal analysis**: A.G., D.E., D.S., J.C.

**Supervision**: A.G., D.E., E. G.

**Funding acquisition**: D.E., E.T.

**Validation**: A.M., C.T., D.E., D.S., J.C.

**Investigation**: D.E, D.S

**Visualisation**: D.E.

**Methodology**: D.E., D.S.

**Project administration**: D.E.

**Writing—original draft**: D.E., D.S.

**Writing—review and editing**: A.G., A.M., C.T, D.E., D.S., E.T, F.V., J.C, J.J.

## Acknowledgments

This work was supported by a 2018 “médecine numérique” (for digital medicine) thesis grant issued by Inserm (French national institute of health and medical research) and Inria (French national research institute for the digital sciences). It was also supported by the Inserm-Inria AI chair EDS-PeriOP and the ANR BrAIN AI chair (ANR-20-CHIA-0016).

## SUPPLEMENTARY MATERIAL

### Supplementary Methods

### EEG processing and feature engineering

We anonymized the monitoring EEG and converted it into the BIDS format^1^ using the MNE-BIDS package^2^. We epoched the signal of each patient using 60-second sliding windows (10 seconds shift) using the MNE-Python software^3^ and the MNE-BIDS pipeline^1^ For every epoch we computed two different types of power-spectral features using the coffeine package^2^. We estimated the power-spectral density (PSD) using Welch’s method and Hamming windows of 8 seconds (4 seconds shift), which can reveal EEG signatures commonly used in anaesthetic monitoring and clinical research. PSDs were estimated in 244 frequency bins between 0 and 30 Hz, and Hamming windows were averaged using a trimmed mean (cut = 25%) to increase robustness to artefacts. To explore the importance of spatio-spectral patterns, we computed the covariance between all 4 electrodes in 5 frequency bands adapted from reference publications on brain-age prediction^4, 5^.

Subsequently, epochs with peak-to-peak amplitude below 0.1 μV on any of the four electrodes were discarded, avoiding inclusion of artefacts or burst suppression. We then selected epochs of stable anaesthesia, defined by the spectral edge frequency 95 (SEF) falling in the [8-13]Hz range^6^. We retained the longest period of consecutive epochs of stable anaesthesia using the average SEF95 across the 4 channels. The power spectral density and the covariance matrices were then averaged across the selected epochs.

### Construction of prediction models, model comparisons and statistical inference

For constructing brain age models, we focused on moderate-risk patients with low ASA scores under propofol anaesthesia (strictly below 3, n = 170, see ds1 in **Table 2**), considering four alternative models highlighting distinct summaries of EEG power spectra. The ‘total power’ model summed EEG power (1-30Hz), averaged across all 4 electrodes (1 feature), representing the hypothesis of spectrally uniform information. The ‘alpha power’ model added EEG power averaged between (8-13Hz) across all 4 electrodes (1 additional feature), representing the hypothesis of a local spectral effect in the frequency range used for monitoring anaesthetic depth. The ‘power spectrum’ model presented uniformly spaced frequencies between (1-30Hz) averaged across all 4 electrodes (16 features), representing the hypothesis of distributed spectral effects. Finally, the ‘spatial patterns’ model implemented the brain age model from a reference publication^4^ which analysed the covariance in 5 frequency bands (low [0.1-1.5]Hz, delta [1.5-4]Hz, theta [4-8]Hz, alpha [8-15]Hz and beta [15-30]Hz), representing the hypothesis of distributed spatio-spectral effects (35 features). The model concatenated the band-specific EEG covariances after vectorization with Riemannian embeddings that accounted for non-linearities caused by EEG-volume conduction^7^.

As in previous reference models^4, 8, 9^, ridge regression^10^ was used as the prediction algorithm. Its regularisation parameter was tuned by generalised cross validation^11^ on a logarithmic grid of 100values between [10^−10^, 10^100^] on each training set. For fair comparisons between models based on different numbers of features, the stacking method^12^ was used as in previous brain-age publications^4, 13^. As the sample size was limited, instead of the non-linear random forest algorithm, we also used ridge regression in the stacking layer.

To estimate the expected generalisation performance, Monte Carlo cross validation (CV) with 100 splits and 20% testing data was applied with fixed random seeds supporting pairwise model comparisons. To date, no generally accepted procedure exists for defining statistically justified null-hypothesis significance tests of observed performance rankings between pairs of prediction models. However, the cross-validation distribution itself provides valuable uncertainty estimates. To gauge practical significance, we followed the strategy from previous work^4, 8^ and plotted or reported percentiles of the CV distribution (P_2.5_, P_25_, P_75_, P_97.5_) alongside rank statistics counting on how many splits one model was better than the other. Chance level was assessed by a dummy regressor predicting the mean outcome of the training data for each sample.

### Detection of burst suppression

To detect iso-electrical suppression (first part of the burst suppression pattern) from intraoperative EEG we adapted the method of a recent publication^14^. For each EEG, a trained clinician identified intra-operative periods based on the alpha-band. From this intraoperative EEG signal, we discarded flat artefacts searching for segments below 0.1 μV and high-amplitude epochs above 80 μV lasting at least 1 second. After the smoothing using a 30-seconds rolling average, we collected regions below 2.5 *μV* amplitude, then we sequentially applied a 0.2 second erosion, a 1 second dilation and an 0.8 second erosion. From this output we estimated the fraction of time spent in burst suppression during induction (the first 25 min) and maintenance periods (after 25 min). We focused on the maintenance period for the better statistical properties of the signal (more samples due to a longer period, less artefacts) and potentially lower false positive rate (drug-induced burst suppression can arise at induction more often). Burst suppression events were automatically excluded from analysis of EEG via artefact rejection. Annotations were not provided for 6 recordings (ds3 - ds5 in **Table 2**).

### Statistical analysis

To investigate correlations between age and the power spectrum, we computed linear mixed effect models using the lm4 package in R^15^. Statistical inference was obtained using confidence intervals and p-values approximated from the t-statistic as implemented by the sjPlot package (https://cran.r-project.org/web/packages/sjPlot/index.html). To investigate the associations between age, brain age and burst suppression across clinical groups, ordinary least squares regression was used on logit-transformed burst-suppression proportions as outcome. This approach allowed us to gauge complementarity of brain age and age in one modelling step^8, 16^. Next, to obtain parameter estimates and p-values, we reported confidence intervals based on the parametric bootstrap implemented in the arm package^17^ as in a previous publication^16^.

## Supplementary Figures

**Fig S1.**
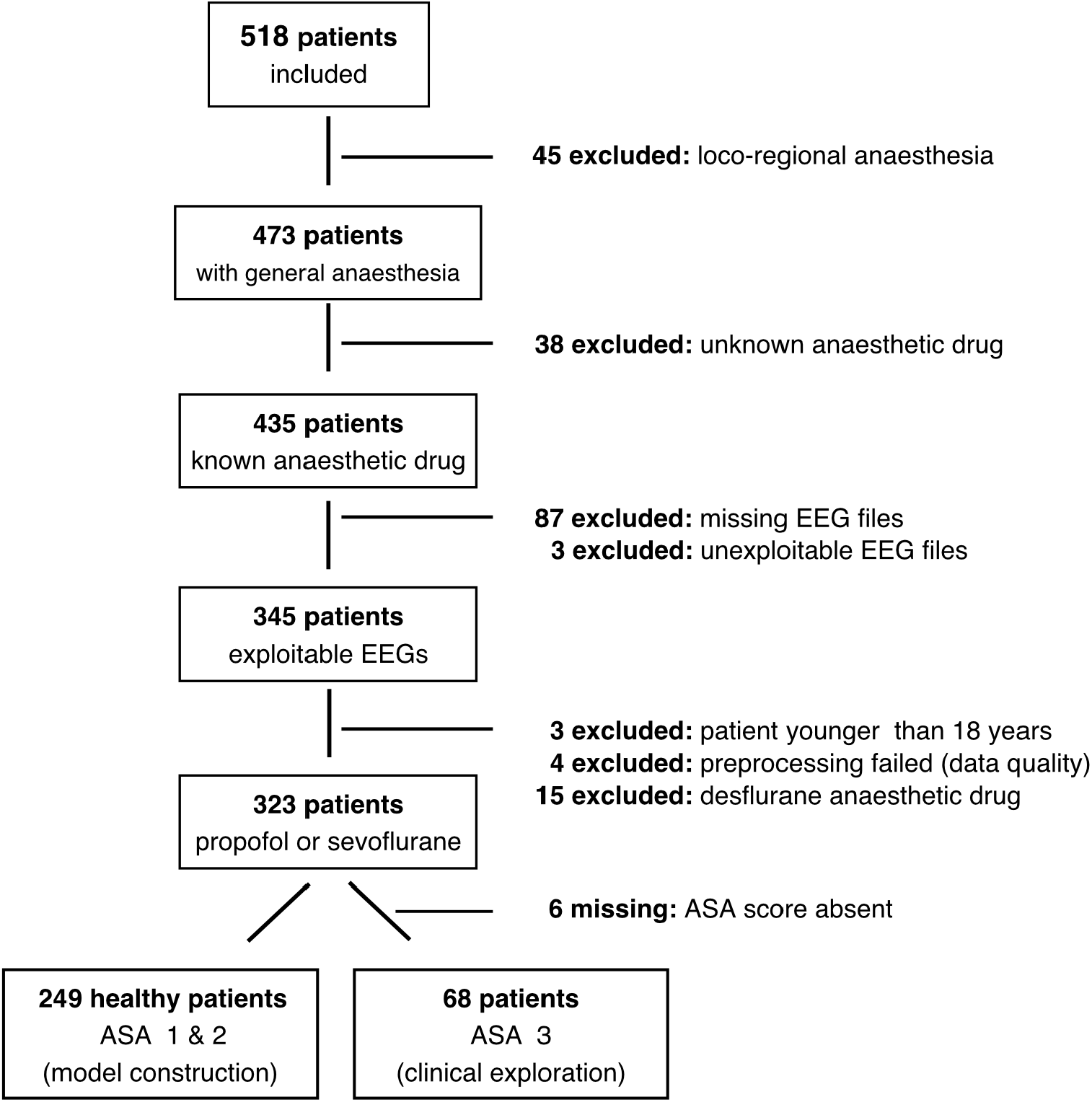
Flowchart illustrating data selection.

**Fig. S2:**
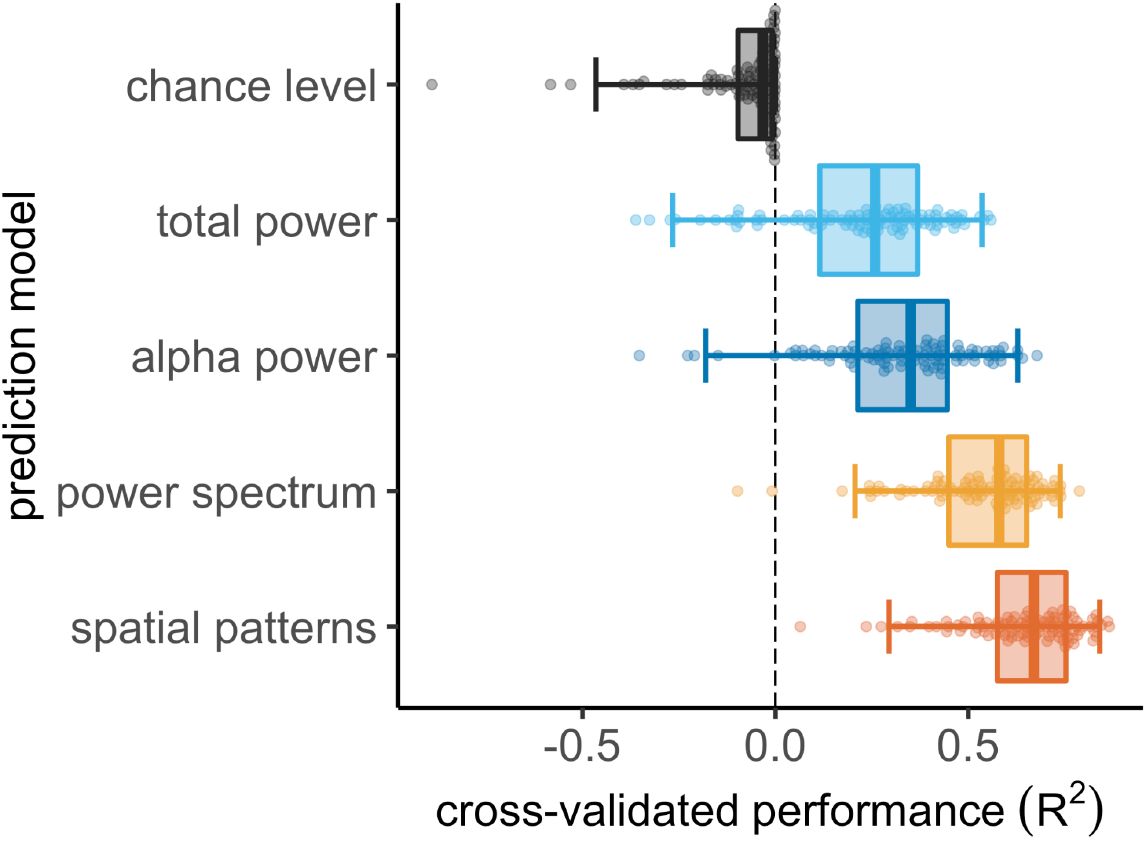
Model comparisons using variance explained (R^2^) as error metric. Same model and visual conventions as in **Fig 2B**.

**Table S1.** Linear Mixed Effect model of EEG log power as a function of frequency and age.

| psd |  |  |  |
| --- | --- | --- | --- |
| *Predictors* | *Estimates* | *CI* | *p* |
| (Intercept) | -30.51 | -31.80 – -29.22 | **<0.001** |
| freqs [log] | -8.62 | -8.77 – -8.48 | **<0.001** |
| age | -0.10 | -0.13 – -0.08 | **<0.001** |
| freqs [log] * age | 0.00 | -0.00 – 0.00 | 0.286 |
| **Random Effects** |  |  |  |
| $\sigma^2$ | 25.73 | | |
| $\tau_{00}$ participant_id | 8.03 | | |
| N participant_id | 166 |  |  |
| Observations | 42496 |  |  |

**Table S2.** Generalised linear model of logit proportion of burst suppression as a function of age and brain age and risk status captured by the ASA score (sc = scaled).

| *Predictors* | **qlogis(y)** |  |  |
| --- | --- | --- | --- |
|  | *Estimates* | *CI* | *p* |
| (Intercept) | -4.31 | -4.59 – -4.04 | **<0.001** |
| age sc | 0.07 | -0.30 – 0.44 | 0.718 |
| brain age sc | 0.86 | 0.47 – 1.25 | **<0.001** |
| risk [high risk] | 0.33 | -0.35 – 1.01 | 0.340 |
| age sc * brain age sc | 0.13 | -0.10 – 0.35 | 0.265 |
| age sc * risk<br>[high risk] | 0.58 | -0.31 – 1.47 | 0.200 |
| brain age sc * risk<br>[high risk] | -0.99 | -1.85 – -0.13 | **0.025** |
| (age sc * brain age sc) *<br>risk [high risk] | 0.34 | -0.25 – 0.93 | 0.254 |
| Observations | 204 |  |  |
| R<sup>2</sup> / R<sup>2</sup> adjusted | 0.349 / 0.326 |  |  |

**Table S3.**
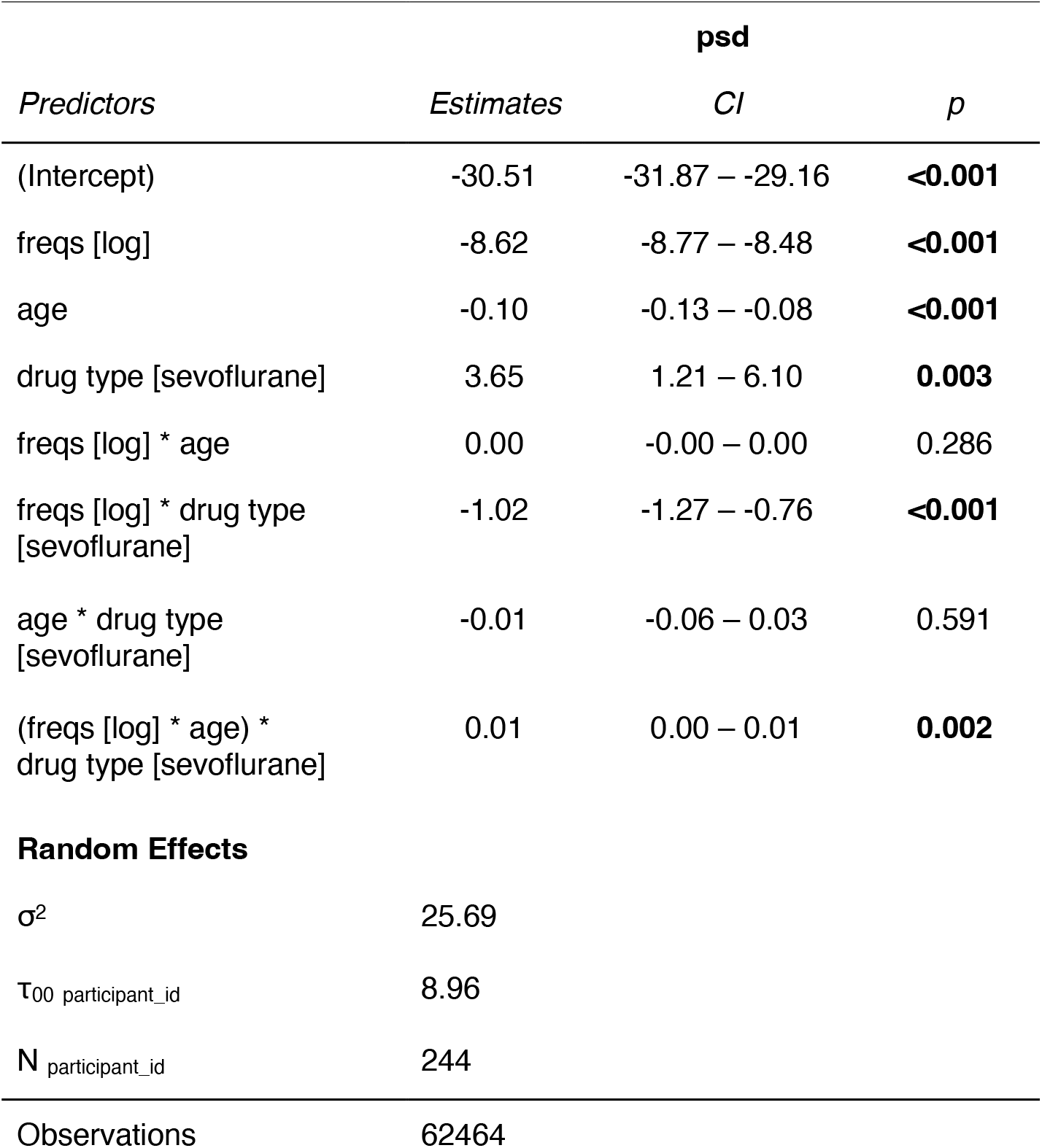
Linear Mixed Effect model of EEG log power as a function of frequency, age and drug type.

## Footnotes

1 https://mne.tools/mne-bids-pipeline/

2 https://github.com/coffeine-labs/coffeine

